# Smoking trajectories and brain health in middle-aged adults: a 15-year longitudinal study

**DOI:** 10.64898/2026.11.13.26368016

**Authors:** Isabel K. Schuurmans, Fleur de Ruijter, Frank J. Wolters, Ryan L. Muetzel, Ingmar H.A. Franken, Julia Neitzel, Janna Cousijn, Hanan El Marroun

**Author notes:** **Correspondence:** Isabel K. Schuurmans, Department of Child and Adolescent Psychiatry and Psychology, Erasmus MC University Medical Center Rotterdam, PO Box 2040, 3000 CA Rotterdam, the Netherlands.

## Abstract

**Background:** Smoking is a prevalent and preventable risk factor for poor brain health. These habits often establish in early adulthood and have lasting consequences, yet most research focuses on later-life smoking, overlooking important prevention windows. We examined smoking trajectories from early adulthood in relation to midlife brain health.

**Methods:** In 1,900 adults (65.6% female, mean age 32.5 years) from the Dutch population-based Generation R Study (ORACLE subset), smoking was assessed five times over 15 years, from before pregnancy to midlife. K-means clustering identified smoking trajectories, alongside measures of current smoking, pack-years, and abstinence years. At mean age 47.7 years, MRI assessed gray and white matter volumes, white-matter hyperintensity volume, cerebrovascular markers, and vertex-wise cortical thickness and surface area.

**Findings:** Five trajectories were identified: non-smoking (63%), quitting before pregnancy (12%), quitting during/after pregnancy (11%), relapsing (9%), and persistent smoking (4%). All compared with non-smoking, quitting before pregnancy was associated with 6.57 cm^3^ larger gray matter (95%CI[0.27,12.87],*p*=.041), 7.71 cm^3^ larger white matter (95%CI[0.77,14.65],*p*=.030), and larger cortical surface area in bilateral middle/inferior temporal regions. Quitting during/after pregnancy was associated with more lacunes (OR=2.39; 95%CI[1.23,4.68],*p*=.011), but similar brain volumes. Relapsing was associated with 0.15 standard deviations more white matter hyperintensity volume (95%CI[0.00,0.30],*p*=.045) and thinner regions in the right cortex. Persistent smoking had double white matter hyperintensities (0.31 standard deviations; 95%CI[0.10,0.50], *p*=.003) and widespread bilateral thinner cortices. Similarly, single-timepoint measures like current smoking (vs never smoking) were linked to 9.72 cm^3^ smaller gray matter volume (95%CI[-16.53,-2.19],*p*=.005), 0.17 standard deviations more hyperintensity volume (95%CI[0.03,0.32],*p*=.015), and widespread thinner cortices. Each additional pack-year was associated with 0.45 cm^3^ smaller gray matter volume (95%CI[-0.85,-0.06],*p*=.025); each additional abstinence year was associated with 0.01 standard deviations less hyperintensity volume (95%CI[-0.02,0.00],*p*=.022).

**Interpretation:** Smoking trajectories from early adulthood and midlife are associated with midlife brain health, supporting early and sustained smoking cessation.

**Funding:** HorizonEurope Research and Innovation Programme (HappyMums, grant agreement No 101057390); European Research Council (TEMPO, grant agreement No 101039672; ORACLE, grant agreement No 678543).

## Introduction

Worldwide, smoking is a significant public health concern. In 2020, an estimated 22.3% of individuals aged above 15 years used tobacco, with the prevalence following an inverted U-shaped pattern across age and peaking at around 50 years.^1^ Smoking cessation is difficult but achievable, and evidence-based interventions can result in sustained abstinence in a substantial proportion of individuals.^2^ Beyond its established detrimental effects on physical and vascular health,^3^ smoking has been linked to poorer mental health, including more depression, anxiety, and cognitive decline.^4^ These findings underscore the importance of understanding how smoking affects the brain.

Previous studies have demonstrated robust associations between smoking and adverse brain outcomes across mid- and late life. In the UK Biobank (>32,000 participants), both current and lifetime smoking were associated with smaller gray and white matter volumes, with dose-response associations with pack-years.^5^ Lifetime smoking has also been associated with greater white matter hyperintensity burden,^6^ higher prevalence of lacunes and cortical infarcts,^7^ and higher prevalence of cerebral microbleeds.^8^

However, several gaps remain. First, smoking is a dynamic behavior rather than a stable trait: quitting commonly requires repeated attempts, and relapse is frequent.^9^ Longitudinal studies have identified distinct adult smoking trajectories, including persistent smoking, quitting, relapse, and decreasing use,^10^ which differ in their associations with lung disease and mortality.^11^ Yet, whether such trajectories relate to brain structure has rarely been examined. These associations are biologically plausible: experimental animal studies indicate that repeated nicotine exposure can induce persistent structural and synaptic plasticity in brain regions involved in reward and behavioral control, which may contribute to the development and maintenance of nicotine dependence.^12^ Second, smoking patterns established and modified earlier in adulthood may have consequences extending into later life, with earlier cessation associated with greater long-term health benefits.^13^ However, much of the literature associating smoking with brain health has focused on smoking measured in mid- or later life, potentially overlooking both the contribution of earlier changes in smoking behavior and opportunities for prevention before exposure cumulates. Third, it remains unclear whether trajectories add information beyond the conventional measures used in most neuroimaging studies, such as ever versus never smoking, which can merge behaviorally distinct groups. In adolescents, trajectory-based approaches have captured meaningful heterogeneity in smoking exposure and may be more informative than simple indicators for predicting later mental health.^14^ Whether trajectory-based modeling adds information for adult brain health remains unclear.

We therefore investigated smoking behavior and brain health in middle-aged adults from a population-based cohort. Smoking was assessed at five time points over approximately 15 years, allowing us to derive longitudinal smoking trajectories. Brain health was subsequently assessed using global brain volumes, cerebrovascular markers, and hypothesis-free vertex-wise measures of cortical thickness and surface area. We aimed to (1) identify smoking trajectories; (2) examine their associations with midlife brain health; and (3) evaluate whether trajectory-based approaches capture associations missed or obscured by single-timepoint measures.

## Methods

### Participants

This study was conducted within the Origins of Alzheimer’s Disease Across the Life Course (ORACLE) Study,^15^ embedded in the population-based Generation R Study.^16^ Generation R enrolled pregnant females and their partners living in Rotterdam, the Netherlands, with expected delivery dates between April 2002 and January 2006. Baseline therefore reflects the period when participants were expecting a child, with intake during the first trimester of pregnancy. Approximately 15 years later, a selected group was invited for an ORACLE follow-up focused on adult brain health. This group comprised Generation R (i) Focus cohort participants, who had undergone extensive earlier assessments; (ii) a random sample of other participants; and (iii) females with a history of hypertensive disorders of pregnancy.

Of 2,083 ORACLE participants, we excluded those missing three or more of five smoking assessments (n=116), those without any usable neuroimaging sequence (n=56), and those with major incidental neuroimaging findings that could distort structural measures (e.g., meningiomas >3 cm; n=11). The final analytical sample included 1,900 participants.

Generation R and ORACLE were approved by the Medical Ethics Committee of Erasmus Medical Center and conducted in accordance with the Declaration of Helsinki. All participants provided written informed consent.

### Smoking variables

Self-reported smoking behavior was assessed using postal questionnaires and operationalized as (1) repeated smoking status and (2) retrospective single-timepoint measures of smoking exposure.

#### Repeated smoking status

Smoking was measured over approximately 15 years. At intake, participants retrospectively reported smoking before pregnancy and current smoking during pregnancy (or until pregnancy was known). Current smoking was subsequently assessed at 6, 10, and 14 years after childbirth. Females reported their own smoking at all waves and their partner’s smoking before pregnancy and at 6 and 14 years postpartum. Males reported their own smoking at intake and 10 years postpartum. Across waves, smoking was harmonized into a binary indicator of current smoking (yes/no); details are provided in **Supplementary Table S1**.

#### Single-timepoint measures of smoking exposure

At 14 years after childbirth, detailed retrospective smoking information was collected, from which we derived three single-timepoint measures: smoking status, pack-years, and abstinence years.

Smoking status was assessed using the question *“Did you ever smoke?”* and categorized into never smoking, quit smoking, and current smoking.

Pack-years were calculated for those who indicated they had quit or were currently smoking at 14 years after childbirth, by multiplying smoking duration (in years) by average packs smoked per day. Smoking duration was calculated as age at smoking cessation minus age at smoking initiation minus any reported periods of temporary cessation for quitting, and as current age minus age at initiation minus temporary cessation when currently smoking. Average packs per day were derived from a categorical question on daily cigarette consumption. Daily cigarette categories were converted to packs using category midpoints (<1/day [0.025 packs], 1-4/day [0.125 packs], 5-9/day [0.35 packs], 10-14/day [0.60 packs], 15-19/day [0.85 packs], and ≥20/day [1 pack]).

Abstinence years were calculated for those who indicated they had quit at 14 years after childbirth and were defined as the number of years since smoking cessation.

### Brain health assessment

Neuroimaging was performed approximately 15 years after inclusion using a 3 Tesla GE Discovery MR750w scanner (General Electric, Milwaukee, WI, USA) with an 8-channel head coil. The scanning protocol and quality-control procedures have been described previously.^15^ Brain health was assessed across three domains: (1) total gray matter, white matter, and white matter hyperintensity volumes; (2) cerebrovascular markers; and (3) hypothesis-free vertex-wise cortical thickness and surface area.

#### Brain volumes

Total gray matter, total white matter, and white matter hyperintensity volumes were derived from T1-weighted images (inversion recovery GRE; 3D resolution 1×1×1 mm voxels; two-fold accelerated ARC). Images were processed using FreeSurfer version 6.0 with default setting,^17^ which provides segmentation of gray matter, white matter, and cerebrospinal fluid. Surface-based models were generated and anatomically labeled using the Desikan-Killiany atlas.^18^ Outcomes from low-quality scans were set to missing and imputed.

#### Cerebrovascular markers

Lacunes and cortical infarcts were assessed on FLAIR images (2D acquisition; voxel size 0.8×1.1×2.5 mm) by trained researchers. Cerebral microbleeds were assessed on T2*-weighted images (3D acquisition; voxel size 0.8×1.1×1 mm; echo time >30 ms), defined as hypointense foci ≤10 mm, and identified through visual inspection by trained raters. Outcomes from low-quality scans were set to missing and imputed.

#### Hypothesis-free, vertex-wise analyses of cortical thickness and surface area

Cortical thickness and surface area were derived from T1-weighted images (inversion recovery GRE; 1×1×1 mm voxels) processed with FreeSurfer version 6.0. Outcomes from low-quality scans were excluded in vertex-wise analyses.

### Covariates

Covariates were assessed at intake by postal self-report questionnaire and included age at neuroimaging, sex, country of origin, educational level, household income, body mass index (BMI), and alcohol use before pregnancy. Age at neuroimaging was calculated from date of birth and imaging date. Country of origin was based on participants’ parents’ countries of birth^16^ and categorized as (i) Europe excluding Turkey, North America, Oceania, and Indonesia, or (ii) Africa, Asia excluding Indonesia, South America, and Turkey. Education was categorized as low (none/primary), intermediate (vocational/higher secondary), or high (Bachelor, Master, or PhD). Household income was dichotomized at €2,200/month, the modal salary in the Netherlands at baseline. Because females were pregnant at baseline, female BMI used retrospectively reported pre-pregnancy weight; male BMI used self-reported weight at intake. Alcohol use before pregnancy was categorized as <1 glass/week, ≥1 glass/week but <1 glass/day, or ≥1 glass/day. For sensitivity analyses, we additionally assessed hypertensive disorders of pregnancy, specifically gestational hypertension and preeclampsia, defined as systolic blood pressure≥140 mmHg or diastolic blood pressure ≥90 mmHg after 20 weeks of gestation, without or with proteinuria, respectively.

### Statistical analysis

Analyses were conducted in R version 4.5.1. White matter hyperintensity volume was natural log-transformed because of skewness and standardized to facilitate interpretation. Missing data were handled using multiple imputation by chained equations with 10 imputed datasets and 10 iterations per dataset; estimates were pooled using Rubin’s rules.^19,20^

#### Smoking trajectories and brain health

Smoking trajectories were identified using k-means clustering of repeated smoking-status measurements. This unsupervised approach iteratively assigns participants to groups without imposing trajectory shapes.^21^ The number of trajectories was selected using (i) the Calinski-Harabasz criterion,^22^ (ii) consistency with prior literature, and (iii) retention of meaningful between-group heterogeneity.

Associations between trajectories and brain health were examined using linear regression for continuous outcomes (gray matter, white matter, and white matter hyperintensity volumes), logistic regression for binary cerebrovascular outcomes (lacunar infarcts, cortical infarcts, and cerebral microbleeds), and vertex-wise analyses for local cortical thickness and surface area. Vertex-wise data were registered to standard stereotaxic space and smoothed with a 10-mm full-width at half-maximum Gaussian kernel, allowing identification of spatially distributed cortical clusters without restricting analyses to predefined regions. The lowest-exposure trajectory (non-smoking) served as reference. Effect estimates are reported as adjusted mean differences for continuous outcomes and odds ratios for binary outcomes.

#### Comparison with single-timepoint smoking measures

Association analyses were repeated using smoking status (never [reference], quit, current) in the full sample, pack-years among former and current smoking participants, and abstinence years among former smoking participants. The same outcome-specific analytical framework was used.

#### Sensitivity analyses

Three sensitivity analyses were restricted to brain volumes (gray matter, white matter and white matter hyperintensity volume), as cerebrovascular markers were too sparse. We tested sex-by-smoking interactions, excluded women with a history of hypertensive disorders of pregnancy, and added a random intercept for family to account for clustering of partners within families.

#### Covariate adjustment

Minimally adjusted models included age at neuroimaging and sex; fully adjusted models additionally included country of origin, education, household income, BMI, and alcohol use before pregnancy. Fully adjusted models were considered the primary analyses. Vertex-wise analyses were conducted only in fully adjusted models.

#### Multiple-test correction

Vertex-wise analyses were cluster-wise corrected for multiple testing.^23^

## Results

### Study population characteristics

The analytical sample included 1,900 participants, of whom 65.6% were female. Mean age was 32.5 years (SD 4.8) at intake and 47.7 years (SD 4.9) at neuroimaging (**Table 1**).

**Table 1.**
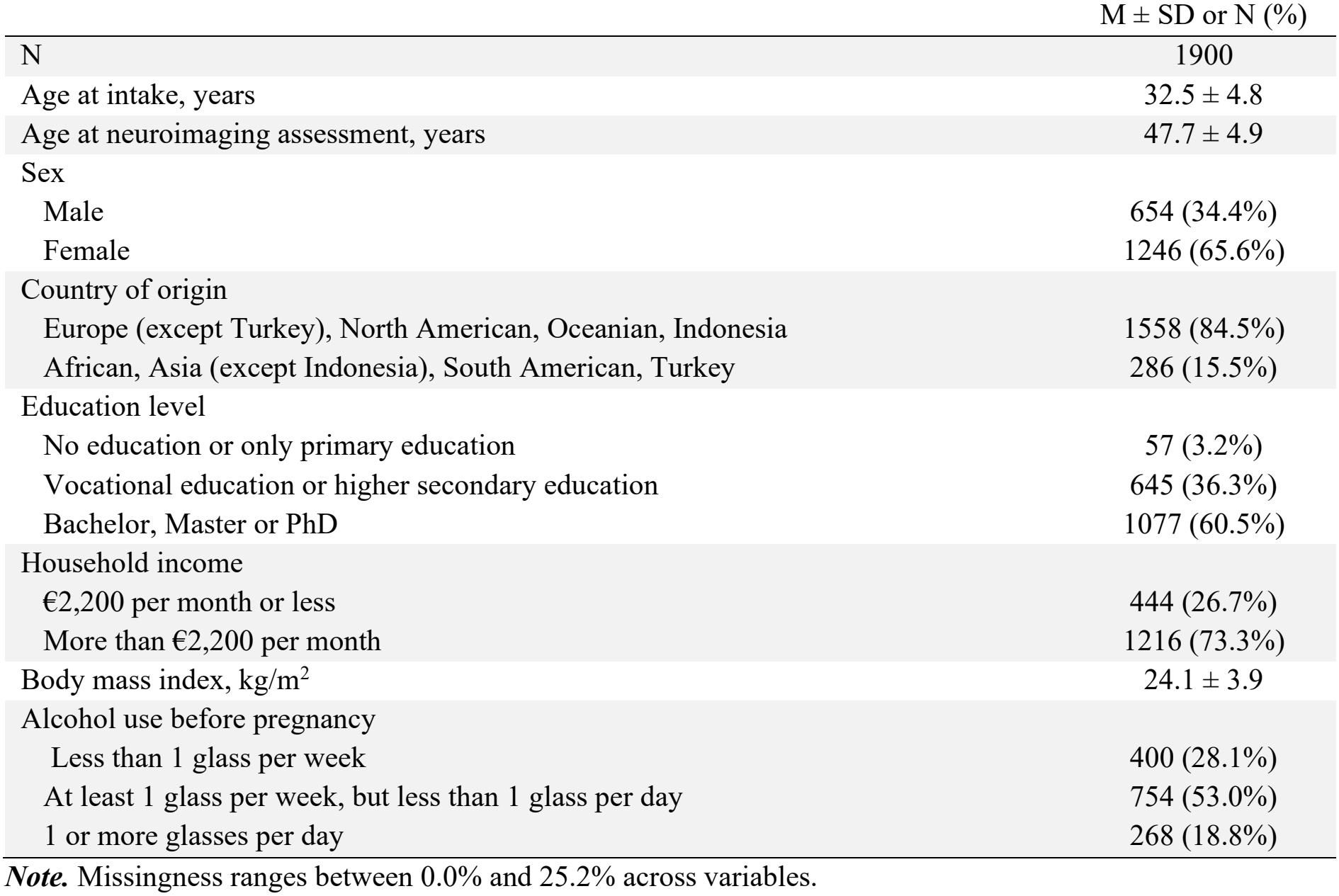
Participant characteristics of the study sample.

### Smoking trajectories and subsequent brain health

The optimal solution comprised five trajectories (**Figure 1A**): non-smoking (n=1,203; 63.3%), quit before pregnancy (n=234; 12.3%), quit during/after pregnancy (n=216; 11.4%), relapsing (n=163; 8.6%), and persistent smoking (n=84; 4.4%). Characteristics by trajectory are shown in **Table S2**. Trajectories characterized by more smoking included fewer highly educated participants and more participants with lower household income.

**Figure 1.**
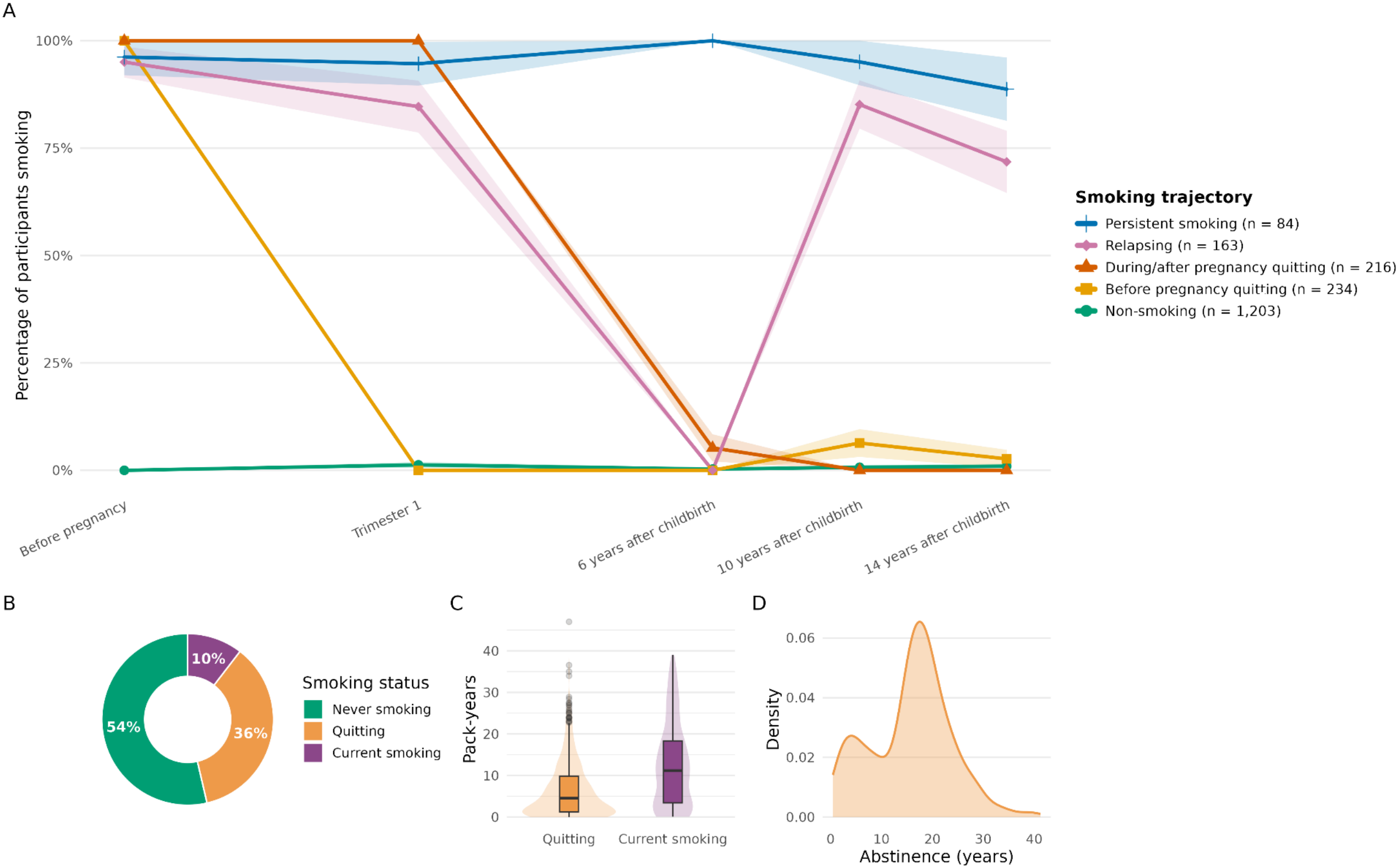
Overview of smoking-related measures. ***Note*. Panel A** shows smoking trajectories over time, with the proportion of individuals within each trajectory who smoked at each time point. **Panel B** shows smoking status at 14 years after childbirth (never smoking, quitting, and current smoking). **Panel C** shows the distribution of pack-years among quitting and current smoking at 14 years after childbirth. **Panel D** shows the distribution of years of abstinence among quitting at 14 years after childbirth.

#### Brain volumes

In fully adjusted models, compared with the non-smoking trajectory, participants who quit before pregnancy had 6.57 cm^3^ greater total gray matter volume (95%CI[0.27,12.87]; *p*=.041) and 7.71 cm^3^ greater total white matter volume (95%CI[0.77,14.65]; *p*=.030), with similar white matter hyperintensity volume (**Figure 2**). Participants who quit during/after pregnancy had similar gray matter, white matter, and white matter hyperintensity volumes to non-smoking. The relapsing trajectory had similar total gray and white matter volumes but 0.15 standard deviations more white matter hyperintensity volume (95%CI[0.00,0.30]; *p*=.045). The persistent smoking trajectory had 0.30 standard deviations more white matter hyperintensity volume (95%CI[0.10,0.50]; *p*=.003).

**Figure 2.**
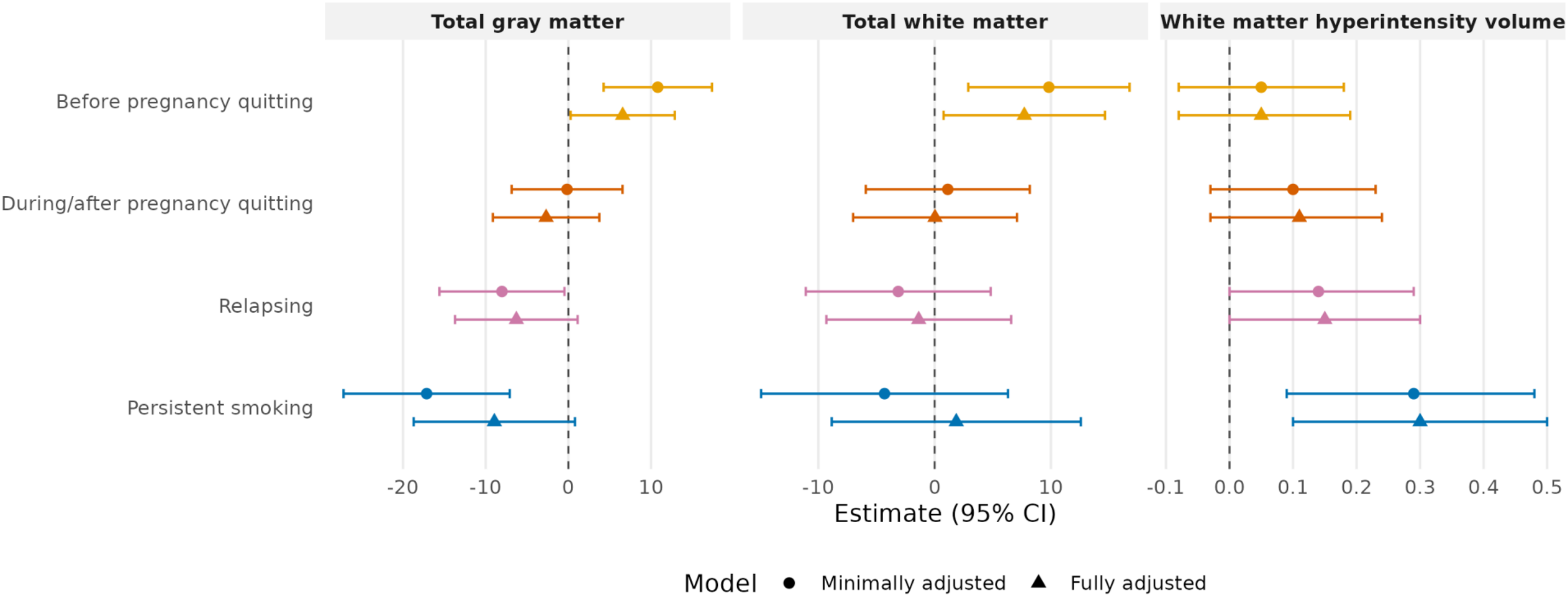
Trajectories of smoking and global neuroimaging measures. ***Note*. N**on-smoking is the reference group. Minimally adjusted models account for age at neuroimaging assessment and sex; Fully adjusted models additionally account for country of origin, education level, household income, BMI, and alcohol use before pregnancy. Estimates for total gray and white matter represent volume cm3. White matter hyperintensity was natural log transformed and standardized, estimates therefore represent standard deviations. Results for lacunes, cortical infarcts, and cerebral microbleeds are in the supplement (Table S3).

#### Cerebrovascular markers

In fully adjusted models, participants in the quitting during/after pregnancy trajectory had 2.39 times the odds of lacunar infarcts compared with non-smoking (95%CI[1.23,4.68], *p*=.011). No other associations were observed for lacunes, cortical infarcts, or cerebral microbleeds (**Table S3**).

#### Vertex-wise cortical thickness and surface area

In fully adjusted analyses (n=1,847), persistent smoking was associated with thinner cortex in bilateral frontal and parietal regions, as well as left cingulate, fusiform/parahippocampal, and supramarginal and right lateral occipital and medial orbitofrontal regions (mean differences 0.07-0.10 mm; **Figure 3A; Table S5**). Relapsing smoking showed more restricted thinning in right frontal, medial orbitofrontal, and paracentral regions (mean differences 0.06–0.08 mm; **Figure 3B; Table S5**). For cortical surface area, quitting before pregnancy was associated with larger bilateral middle and inferior temporal surface area (mean difference 0.04 mm^2^ in both hemispheres; **Figure 3C; Table S6**).

**Figure 3.**
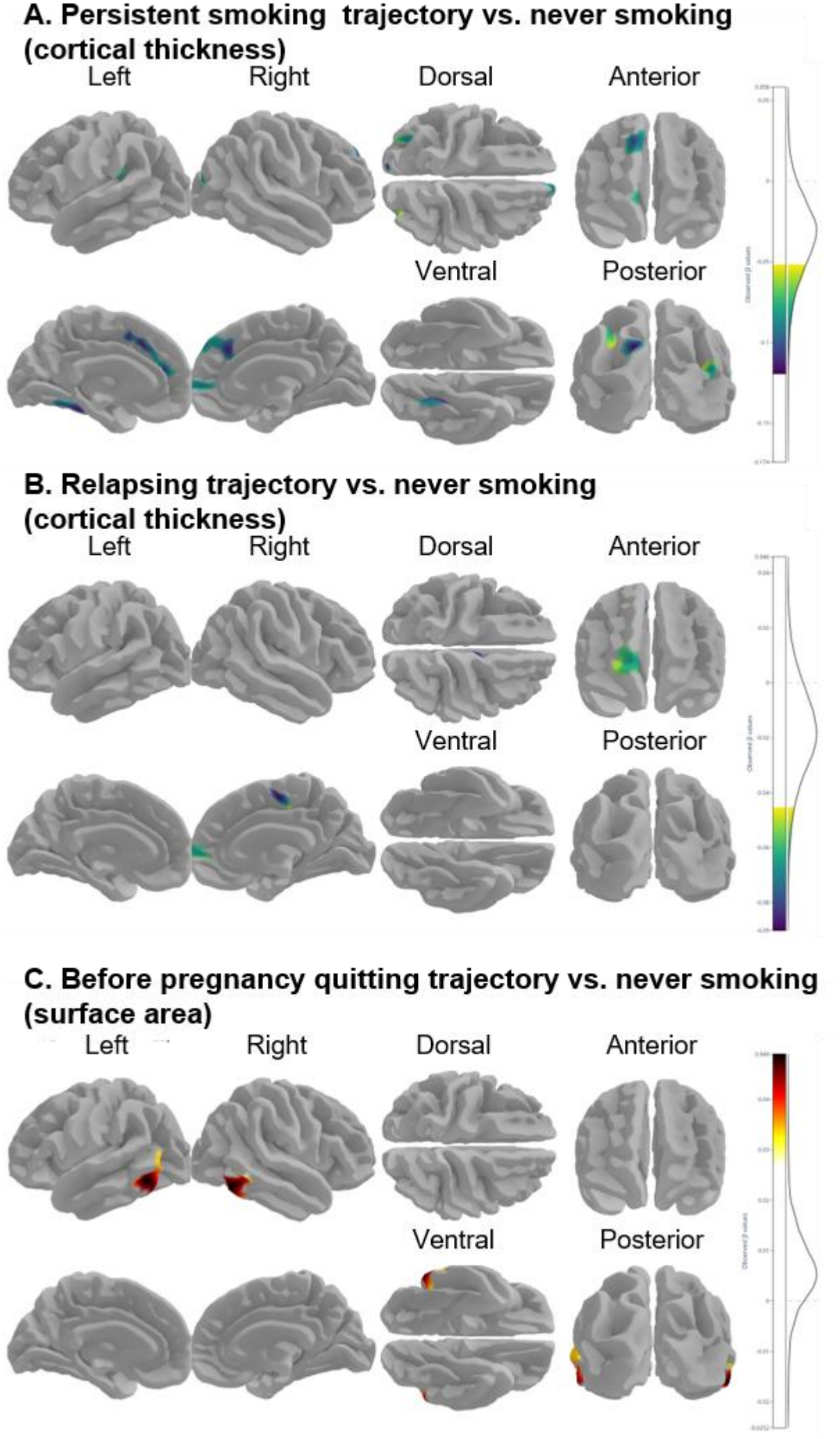
Associations between smoking trajectories and regional measures of cortical thickness and surface area. ***Note***. These are results from the fully adjusted model, which accounts for age at neuroimaging assessment, sex, country of origin, education level, household income, BMI, and alcohol use before pregnancy. Vertex-wise analyses were cluster-wise corrected for multiple testing. Visualization was done with verywise (https://seredef.github.io/verywise/).

### Single-timepoint markers of smoking and subsequent brain health

During the 14-years after childbirth assessment, 54% of participants had never smoked, 36% had quit, and 10% currently smoked (**Figure 1B**). Among participants with a smoking history (n=831), mean pack-years were 8.0 (SD 8.2), with lower exposure among former than current smoking participants (6.6 [SD 7.0] vs 12.5 [SD 10.1]; **Figure 1C**). Among former smoking participants (n=644), mean abstinence duration was 15.9 years (SD 7.9; **Figure 1D**).

#### Volume of total white matter, gray matter, and white matter hyperintensities

Results are presented in **Table 2**. In fully adjusted models and compared with never smoking, current smoking was associated with 9.72 cm^3^ smaller total gray matter volume (95%CI[-16.53,-2.91], *p*=.005) and 0.17 standard deviations more white matter hyperintensity volume (95%CI[0.03,0.32], *p*=.015), but not with white matter volume. In contrast, participants who quit smoking had similar volumes to those who never smoked. Yet, among those with a history of smoking, each additional pack-year was associated with 0.45 cm^3^ smaller total gray matter volume (95%CI[-0.85,-0.06], *p*=.025). Among those who quit smoking, each additional year of abstinence was associated with 0.01 standard deviations less white matter hyperintensity volume per year (95%CI[-0.02,0.00], *p*=.022).

**Table 2.**
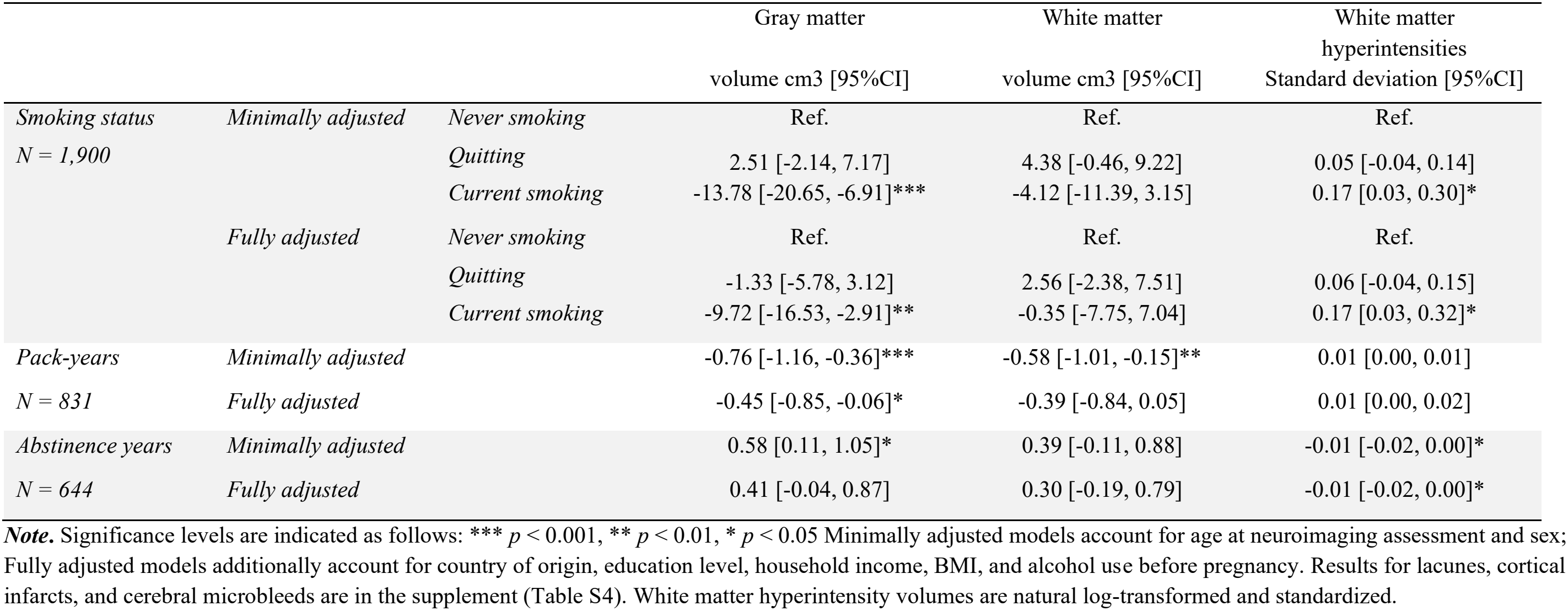
Associations between single-timepoint markers of smoking and global neuroimaging measures.

|  |  |  | Gray matter<br>volume cm3 [95%CI] | White matter<br>volume cm3 [95%CI] | White matter hyperintensities<br>Standard deviation [95%CI] |
|---|---|---|---|---|---|
| *Smoking status* | *Minimally adjusted* | *Never smoking* | Ref. | Ref. | Ref. |
| *N = 1,900* |  | *Quitting* | 2.51 [-2.14, 7.17] | 4.38 [-0.46, 9.22] | 0.05 [-0.04, 0.14] |
|  |  | *Current smoking* | -13.78 [-20.65, -6.91]*** | -4.12 [-11.39, 3.15] | 0.17 [0.03, 0.30]* |
|  | *Fully adjusted* | *Never smoking* | Ref. | Ref. | Ref. |
|  |  | *Quitting* | -1.33 [-5.78, 3.12] | 2.56 [-2.38, 7.51] | 0.06 [-0.04, 0.15] |
|  |  | *Current smoking* | -9.72 [-16.53, -2.91]** | -0.35 [-7.75, 7.04] | 0.17 [0.03, 0.32]* |
| *Pack-years* | *Minimally adjusted* |  | -0.76 [-1.16, -0.36]*** | -0.58 [-1.01, -0.15]** | 0.01 [0.00, 0.01] |
| *N = 831* | *Fully adjusted* |  | -0.45 [-0.85, -0.06]* | -0.39 [-0.84, 0.05] | 0.01 [0.00, 0.02] |
| *Abstinence years* | *Minimally adjusted* |  | 0.58 [0.11, 1.05]* | 0.39 [-0.11, 0.88] | -0.01 [-0.02, 0.00]* |
| *N = 644* | *Fully adjusted* |  | 0.41 [-0.04, 0.87] | 0.30 [-0.19, 0.79] | -0.01 [-0.02, 0.00]* |

***Note.*** Significance levels are indicated as follows: \*\*\* *p* < 0.001, \*\* *p* < 0.01, \* *p* < 0.05 Minimally adjusted models account for age at neuroimaging assessment and sex; Fully adjusted models additionally account for country of origin, education level, household income, BMI, and alcohol use before pregnancy. Results for lacunes, cortical infarcts, and cerebral microbleeds are in the supplement (Table S4). White matter hyperintensity volumes are natural log-transformed and standardized.

#### Cerebrovascular markers

No associations were observed for cortical infarcts, lacunar infarcts, or cerebral microbleeds (**Table S4**).

#### Vertex-wise cortical thickness and surface area

Compared with never smoking, current smoking was associated with widespread cortical thinning across both hemispheres (**Table S5; Figure S2A**, n=1,847), involving bilateral frontal, cingulate, temporal, and paracentral regions, as well as left fusiform, supramarginal, and parietal and right orbitofrontal, pre-/postcentral, and insular regions (mean differences 0.04–0.10 mm). For cortical surface area, higher pack-years were associated with smaller surface area in the left fusiform and inferior temporal regions (mean difference -0.002 mm^2^ per pack-year; **Table S6, Figure S2B**, n=807).

### Sensitivity analyses

Results were largely consistent across sexes, although the association between quitting during/after pregnancy and white matter volume was more negative in females than in males (-16.60 cm^3^, 95%CI[-30.44,-2.76], p=.019), without a significant main effect. After excluding women with hypertensive disorders of pregnancy (11.6%), effect sizes were similar, but associations of before-pregnancy quitting with white matter and of relapsing and current smoking with white matter hyperintensity volume were less or no longer significant, likely reflecting reduced power. Accounting for family clustering did not change results; only relapsing smoking lost significance for white matter hyperintensity volume, with an unchanged estimate.

## Discussion

In this population-based study of middle-aged adults, smoking trajectories across approximately 15 years of prospective measurements were differentially associated with brain health. We highlight three findings. First, repeated smoking assessments identified substantial heterogeneity beyond conventional never, former, and current smoking categories, distinguishing former smoking by the timing of cessation and current smoking by patterns of relapse versus continued smoking. Second, trajectories involving cessation generally showed more favorable brain-health profiles than continued smoking, consistent with the association between longer abstinence and lower white matter hyperintensity burden. Third, trajectories reflecting greater cumulative smoking exposure were associated with less favorable brain health, including greater white matter hyperintensity burden and widespread thinner cortices, a pattern further supported by dose-response associations between pack-years and brain health outcomes.

Our findings underscore that smoking patterns are more heterogeneous than can be captured with classifications of never, former, and current smoking. The repeated data separated participants who stopped smoking before pregnancy from those who stopped later, and distinguished relapsing from persistently smoking participants despite both groups contributing to the broad category of ever or current smoking at later assessments; broadly resembling trajectories identified in a previous general-population study of >3,000 adults followed for 25 years.^10^ Differences from other longitudinal studies likely reflect variation in cohort composition, historical changes in smoking policy and social norms, developmental stage, and follow-up duration.^14,24^ For example, studies beginning in adolescence more often capture smoking initiation and experimentation,^25^ whereas our prospective assessments assessed specifically pregnancy, a period when cessation and relapse are particularly common, with retrospective information on smoking before pregnancy which allowed us to distinguish earlier quitting patterns.

Examining these smoking patterns in relation to brain health showed that the level of detail in smoking history mattered. When former smoking was treated as one category, former smoking participants had similar total gray matter, white matter, and white matter hyperintensity volumes to never-smoking participants. Trajectory analyses, however, separated cessation by timing and revealed meaningful heterogeneity. Those who quit before pregnancy had larger total brain volume and biliteral temporal surface area than non-smoking participants, whereas those who quit during/after pregnancy had similar volumes. Longer abstinence was associated with less white matter hyperintensity volume. These findings are broadly similar to prior evidence of partial brain recovery after smoking cessation,^26^ which could suggest that smoking accelerates age-related volume loss via microvascular damage or direct neurotoxicity (e.g., oxidative stress, reduced oxygen from carbon monoxide, mitochondrial dysfunction), while after smoking cessation, surviving neurons may undergo temporary compensatory structural changes through neuroplasticity, potentially contributing to partial recovery of brain volume. Yet, recovery would not explain the *larger* volumes we observed, pointing towards residual confounding explaining these effects: participants who quit before pregnancy had more favorable socioeconomic profiles, which may suggest that part of the observed associations may reflect broader health and lifestyle differences. Notably, for cerebrovascular outcomes, the trajectory analysis suggested higher odds of lacunar infarcts among participants who had quit during/after pregnancy, although this finding should be interpreted cautiously given the sparse binary outcomes and the absence of a broader pattern across cerebrovascular markers. More generally, because the timing of infarcts relative to smoking cessation is unknown, reverse causation or confounding by indication remains possible if vascular events prompted cessation.^27^ Repeated brain imaging would be required to establish the temporal sequence more clearly.

Greater smoking exposure showed a consistent adverse pattern. Each additional pack-year was associated with smaller gray matter volume, while relapsing and persistent trajectories had greater white matter hyperintensity burden than non-smoking, with the largest difference for persistent smoking. Vertex-wise analyses further differentiated these trajectories. Persistent smoking was associated with widespread thinner cortex across frontal, parietal, temporal, occipital and cingulate regions, whereas relapsing smoking showed a restricted pattern in right superior frontal, rostral middle frontal, medial orbitofrontal, and paracentral regions. The right paracentral lobule has shown withdrawal-related changes in task response, particularly increased response variability during placebo/abstinence conditions,^28^ although the functional implications of regional patterns remain uncertain. Notably, there were substantially fewer associations with surface area, suggesting that cortical thickness may be the more sensitive brain marker of smoking exposure in midlife.

Strengths include prospective and repeated smoking assessments over 15 years together with detailed MRI phenotyping. Assessments around pregnancy provided a natural point at which smoking patterns often change: smoking prevalence and intensity decline during pregnancy because of fetal-health concerns,^29^ after which some individuals maintain abstinence while others relapse. This offered meaningful heterogeneity in cessation timing and subsequent smoking behavior. The combination of global brain volumes, cerebrovascular markers, and hypothesis-free vertex-wise cortical analyses also allowed us to examine both global and spatially localized brain differences. Several limitations should be considered. First, the sample was predominantly female (>65%) and included couples. Because smoking and second-hand smoke exposure may be correlated within couples,^30^ observations may not be independent. Adding a random effect for family in brain volume analyses did not,, change any results. Second, pre-pregnancy smoking was reported retrospectively, introducing recall bias. Moreover, questionnaire data might be susceptible to information bias due particularly to smoking individuals providing socially desirable answers about smoking in relation to pregnancy. This may have led to an underestimation of associations with continued smoking, while associations attributed to quitting may partly reflect the effects of continued smoking among participants who reported quitting but had continued to smoke. Third, midlife follow-up enabled brain health assessment before neurodegenerative disease onset, which reduces reverse causation. Yet, the timing of white matter hyperintensities, infarcts, and microbleeds relative to smoking trajectories remains unknown.

In conclusion, smoking trajectories across approximately 15 years were differentially associated with midlife brain health. Repeated assessments distinguished sustained cessation, later cessation, relapse, and persistent smoking patterns that were obscured by conventional single-timepoint categories. Greater and more persistent smoking

exposure was associated with higher white matter hyperintensity burden, smaller gray matter volume in dose-response analyses, and widespread cortical thinning, whereas sustained cessation generally showed more favorable profiles. These findings indicate early- and mid-adult smoking behavior may shape cumulative exposure long before age-related brain disease becomes clinically apparent, creating a prevention window that is missed when studies assess smoking only near the time of neuroimaging. Findings also support the importance of early and sustained smoking cessation.

### Research in context

#### Evidence before this study

Prior to this study, several population-based studies examined the relationship between smoking and brain health, amongst others focusing on total gray and white matter volume, white matter hyperintensity volume, and cerebrovascular markers. Notable studies include large cohort analyses from the UK Biobank, which showed associations between both current and lifetime smoking and reduced brain volume, particularly gray and white matter, as well as increased white matter hyperintensity volume and higher incidence of cerebral microbleeds. Research on smoking trajectories, however, remains limited, with few studies exploring how long-term smoking behavior, including repeated quit attempts and relapses, is associated with brain health. Furthermore, many studies relied on cross-sectional measures of smoking, overlooking its dynamic and changing nature over time, particularly across life stages such as pregnancy and post-pregnancy where individuals often reconsider smoking behaviors. Earlier studies also mostly categorized smoking behavior into broad groups (never, former, current smoking individuals), which likely missed important variations within smoking trajectories that might relate differently to brain health outcomes.

#### Added value of this study

This study offers new insights by identifying and characterizing five distinct smoking trajectories (i.e., non-smoking, quitting before pregnancy, quitting during/after pregnancy, relapsing, and persistent smoking) over a 15-year period in a large cohort of middle-aged adults. By analyzing smoking behavior across multiple time points, we demonstrated that smoking trajectories are associated with nuanced differences in brain health, revealing a more detailed understanding of how smoking cessation timing and relapse influence neuroanatomy. Key findings include that quitting smoking before pregnancy was linked to larger gray and white matter volumes, while quitting during/after pregnancy was associated with higher odds of lacunes. Persistent smoking was associated with widespread thinner cortices, and relapsing smoking exhibited localized thinner cortices in the right hemisphere.

#### Implications of all the available evidence

These findings highlight that smoking behavior is not a static trait, and the timing of cessation plays a potential role in subsequent brain health. This study also underscores the importance of sustained smoking cessation in promoting better brain health, with more favorable outcomes for those who quit earlier. This could influence public health strategies and interventions, highlighting the importance of early cessation and the need for ongoing support to prevent relapse.

## Data Availability

All data produced in the present study are available upon reasonable request to Generation R Study group board

## Acknowledgements and funding

The general design of the Generation R Study is funded by the Erasmus Medical Center, Rotterdam, ZonMw, the Netherlands Organization for Scientific Research (NWO), and the Ministry of Health, Welfare and Sport, and is conducted by the Erasmus Medical Center in close collaboration with the Faculty of Social Sciences of the Erasmus University Rotterdam, and the Stichting Trombosedienst & Artsenlaboratorium Rijnmond (STAR-MDC), Rotterdam. This project received funding from the European Research Council (ERC) under the European Union’s Horizon 2020 research and innovation programme (project: ORACLE, grant agreement No: 678543).

The work of Isabel Schuurmans and Hanan El Marroun is supported by the European Union’s HorizonEurope Research and Innovation Programme (HappyMums, grant agreement No 101057390). The work of Isabel Schuurmans is additionally supported by the European Research Council (TEMPO; grant agreement No 101039672).

The funders had no role in the study design, data collection, analysis, interpretation of the data, or writing of the report.

## Notes

### Competing Interest Statement

The authors have declared no competing interest.

### Author Declarations

Ethics committee of Erasmus Medical Center a gave ethical approval for this work

