## Supplementary for "Smoking trajectories and brain health in middle-aged adults: a 15-year longitudinal study"

### Supplement

**Table S1. Measurements used to construct smoking trajectories**

| | Mother | | | Partner | | |
|---|---|---|---|---|---|---|
| Timepoint | Reporter | Question | Reclassification | Reporter | Question | Reclassification |
| Before pregnancy | Mother | Did you smoke before pregnancy<br>*0 = No*<br>*1 = Yes* | **No smoking** = 'No'<br>**Smoking =** 'Yes' | Mother | Did the biological father smoke before pregnancy<br>*0 = No*<br>*1 = Yes*<br>*2 = Do not know* | **No smoking** = 'No'<br>**Smoking =** 'Yes'<br>**NA** = 'Do not know' |
| Intake (first trimester) | Mother | Have you smoked in the past 3 months<br>*1 = No*<br>*2 = Yes until I knew*<br>*3 = Yes still* | **No smoking** = 'No'<br>**Smoking =** 'Yes until I knew', 'Yes still' | Partner | Smoking in the 2 months before the pregnancy<br>*0 = No*<br>*1 = Yes* | **No smoking** = 'No'<br>**Smoking =** 'Yes' |
| 6 years postpartum | Mother | Does the child's mother smoke at home?<br>*0 = No*<br>*1 = Yes* | **No smoking** = 'No'<br>**Smoking =** 'Yes' | Mother | Does the child's father smoke at home?<br>*0 = No*<br>*1 = Yes* | **No smoking** = 'No'<br>**Smoking =** 'Yes' |
| 10 years postpartum | Mother | Have you ever smoked?<br>*1 = No, I have never smoked*<br>*2 = Yes, but I do not smoke anymore*<br>*3 = Yes, I still smoke now* | **No smoking** = 'No' & 'Yes, but I do not smoke anymore'<br>**Smoking =** 'Yes, I still smoke now' | Partner | Have you ever smoked?<br>*1 = No, I have never smoked*<br>*2 = Yes, but I do not smoke anymore*<br>*3 = Yes, I still smoke now* | **No smoking** = 'No' & 'Yes, but I do not smoke anymore'<br>**Smoking =** 'Yes, I still smoke now' |
| 14 years postpartum | Mother | Has mother ever smoked?<br>*1 = No, never smoked*<br>*2 = Yes, but no longer smokes*<br>*3 = Yes, still smokes* | **No smoking** = 'No' & 'Yes, but no longer smokes'<br>**Smoking =** 'Yes, still smokes' | Mother | Has he/she ever smoked?<br>*1 = No, never smoked*<br>*2 = Yes, but no longer smokes*<br>*3 = Yes, still smokes* | **No smoking** = 'No' & 'Yes, but no longer smokes'<br>**Smoking =** 'Yes, still smokes' |

*Note*. All questions were completed by the participant at home using a postal questionnaire.

**Table S2. Participant characteristics of the study sample at intake**

| M ± SD or N (%) | Non-smoking | Before-pregnancy quitting | During/after pregnancy quitting | Relapsing | Persistent smoking |
|---|---|---|---|---|---|
| N | 1203 (63.3%) | 234 (12.3%) | 216 (11.4%) | 163 (8.6%) | 84 (4.4%) |
| Age at intake, years | 32.5 ± 4.8 | 32.3 ± 4.0 | 33.0 ± 4.7 | 32.2 ± 4.6 | 31.7 ± 6.7 |
| Age at neuroimaging assessment, years | 47.7 ± 4.9 | 47.4 ± 4.1 | 48.1 ± 4.8 | 47.4 ± 4.8 | 47.4 ± 6.7 |
| Sex | | | | | |
| Male | 406 (33.7%) | 18 (7.7%) | 120 (55.6%) | 76 (46.6%) | 34 (40.5%) |
| Female | 797 (66.3%) | 216 (92.3%) | 96 (44.4%) | 87 (53.4%) | 50 (59.5%) |
| Country of origin | | | | | |
| Europe (- Turkey), North American, Oceanian, Indonesia | 953 (82.1%) | 210 (90.1%) | 199 (93.4%) | 133 (84.7%) | 63 (78.8%) |
| African, Asia (- Indonesia), South American, Turkey | 208 (17.9%) | 23 (9.9%) | 14 (6.6%) | 24 (15.3%) | 17 (21.2%) |
| Education level | | | | | |
| No education or only primary education | 34 (3.0%) | 2 (0.9%) | 5 (2.5%) | 8 (5.4%) | 8 (10.1%) |
| Vocational education or higher secondary education | 378 (33.8%) | 71 (30.6%) | 77 (38.1%) | 68 (45.9%) | 51 (64.6%) |
| Bachelor, Master or PhD | 706 (63.1%) | 159 (68.5%) | 120 (59.4%) | 72 (48.6%) | 20 (25.3%) |
| Household income | | | | | |
| €2,200 per month or less | 267 (25.7%) | 51 (23.8%) | 42 (21.2%) | 45 (32.1%) | 39 (54.9%) |
| More than €2,200 per month | 770 (74.3%) | 163 (76.2%) | 156 (78.8%) | 95 (67.9%) | 32 (45.1%) |
| Body mass index, kg/m$^2$ | 24.1 ± 3.9 | 23.5 ± 4.1 | 24.1 ± 3.8 | 24.5 ± 3.2 | 24.6 ± 4.6 |
| Alcohol use before pregnancy | | | | | |
| Less than 1 glass per week | 280 (33.5%) | 40 (19.4%) | 37 (19.3%) | 27 (21.1%) | 16 (27.1%) |
| At least 1 glass per week but less than 1 glass per day | 446 (53.3%) | 130 (63.1%) | 93 (48.4%) | 57 (44.5%) | 28 (47.5%) |
| 1 or more glasses per day | 111 (13.3%) | 36 (17.5%) | 62 (32.3%) | 44 (34.4%) | 15 (25.4%) |

***Note.*** Missingness ranges between 0.0% and 25.2% across variables.

**Table S3. Associations between smoking-trajectories and brain outcomes**

| | | | Lacunes<br>Odds ratio [95%CI] | Cortical infarcts<br>Odds ratio [95%CI] | Cerebral microbleeds<br>Odds ratio [95%CI] |
|---|---|---|---|---|---|
| *Smoking trajectories*<br>*N = 1,900* | *Minimally adjusted* | *Non-smoking* | Ref. | Ref. | Ref. |
| | | *Before pregnancy quitting* | 0.93 [0.40, 2.15] | 4.30 [1.14, 16.22]* | 1.25 [0.78, 1.99] |
| | | *During/after-pregnancy quitting* | 2.22 [1.16, 4.27]* | - | 1.12 [0.70, 1.80] |
| | | *Relapsing* | 0.60 [0.18, 1.99] | 2.98 [0.75, 11.81] | 0.67 [0.34, 1.31] |
| | | *Persistent smoking* | 0.66 [0.15, 2.84] | - | 1.77 [0.94, 3.33] |
| | *Fully adjusted* | *Non-smoking* | Ref. | Ref. | Ref. |
| | | *Before pregnancy quitting* | 0.96 [0.41, 2.25] | 3.60 [0.93, 13.99] | 1.22 [0.76, 1.95] |
| | | *During/after-pregnancy quitting* | 2.39 [1.23, 4.68]* | - | 1.08 [0.67, 1.74] |
| | | *Relapsing* | 0.60 [0.18, 2.05] | 2.21 [0.51, 9.58] | 0.65 [0.33, 1.28] |
| | | *Persistent smoking* | 0.72 [0.16, 3.22] | - | 1.69 [0.89, 3.24] |

***Note.*** Significance levels are indicated as follows: *** $p < 0.001$, ** $p < 0.01$, * $p < 0.05$. Minimally adjusted models account for age at neuroimaging assessment and sex; Fully adjusted models additionally account for country of origin, education level, household income, BMI, and alcohol use before pregnancy. The non-smoking are the reference group. Estimates for cortical infarcts could not be obtained within the groups of those who quit smoking during/after pregnancy and those who were persistently smoking, as there were no participants in these categories with a cortical infarct.

**Table S4. Associations between smoking and brain outcomes**

| | | | Lacunes<br>Odds ratio [95%CI] | Cortical infarcts<br>Odds ratio [95%CI] | Cerebral microbleeds<br>Odds ratio [95%CI] |
|---|---|---|---|---|---|
| *Smoking status* | *Minimally adjusted* | *Never smoking* | Ref. | Ref. | Ref. |
| *N = 1,900* | | *Quitting* | 1.21 [0.72, 2.05] | 2.58 [0.74, 8.94] | 0.98 [0.70, 1.36] |
| | | *Current smoking* | 0.56 [0.19, 1.62] | 3.43 [0.75, 15.68] | 1.06 [0.65, 1.73] |
| | *Fully adjusted* | *Never smoking* | Ref. | Ref. | Ref. |
| | | *Quitting* | 1.23 [0.71, 2.14] | 2.23 [0.62, 8.05] | 0.94 [0.67, 1.32] |
| | | *Current smoking* | 0.60 [0.20, 1.84] | 2.78 [0.55, 14.02] | 1.00 [0.59, 1.67] |
| *Pack-years* | *Minimally adjusted* | | 1.02 [0.98, 1.06] | 1.02 [0.95, 1.09] | 1.00 [0.97, 1.03] |
| *N = 831* | *Fully adjusted* | | 1.04 [0.99, 1.08] | 1.02 [0.94, 1.10] | 1.00 [0.97, 1.03] |
| *Abstinence years* | *Minimally adjusted* | | 0.98 [0.93, 1.03] | 0.98 [0.89, 1.07] | 1.02 [0.99, 1.06] |
| *N = 644* | *Fully adjusted* | | 0.98 [0.93, 1.03] | 0.99 [0.90, 1.09] | 1.02 [0.99, 1.06] |

***Note.*** Significance levels are indicated as follows: *** $p < 0.001$, ** $p < 0.01$, * $p < 0.05$. Minimally adjusted models account for age at neuroimaging assessment and sex; Fully adjusted models additionally account for country of origin, education level, household income, BMI, and alcohol use before pregnancy.

**Table S5. Vertex-wise analyses full results for cortical thickness**

| Variable | Levels | Regions | Number of vertices | Mean thickness | Thickness mm [95% CI] |
|---|---|---|---|---|---|
| Smoking trajectories | Persistent smoking vs non-smoking | LH superior frontal & rostral anterior cingulate | 1112 | 2.44 | −0.09 [−0.14, −0.04] |
| | | LH inferior parietal & superior parietal | 611 | 2.44 | −0.07 [−0.11, −0.03] |
| | | LH fusiform & parahippocampal | 671 | 2.45 | −0.09 [−0.14, −0.05] |
| | | LH supramarginal | 411 | 2.44 | −0.08 [−0.12, −0.04] |
| | | LH superior parietal | 236 | 2.49 | −0.10 [−0.14, −0.05] |
| | | RH superior frontal | 979 | 2.41 | −0.09 [−0.14, −0.04] |
| | | RH superior frontal & medial orbitofrontal | 297 | 2.48 | −0.09 [−0.13, −0.04] |
| | | RH inferior parietal & lateral occipital | 301 | 2.58 | −0.07 [−0.11, −0.04] |
| | Relapsing vs non-smoking | RH superior frontal, rostral middle frontal & medial orbitofrontal | 494 | 2.42 | −0.06 [−0.09, −0.03] |
| | | RH paracentral & superior frontal | 351 | 2.41 | −0.07 [−0.12, −0.03] |
| Single time-point marker of smoking | Current vs never smoking | LH superior frontal, caudal anterior cingulate & rostral anterior cingulate | 4360 | 2.44 | −0.07 [−0.10, −0.03] |
| | | LH inferior temporal & middle temporal | 427 | 2.43 | −0.07 [−0.11, −0.04] |
| | | LH supramarginal & superior temporal | 577 | 2.46 | −0.05 [−0.08, −0.02] |
| | | LH rostral middle frontal | 298 | 2.44 | −0.06 [−0.09, −0.03] |
| | | LH superior parietal & inferior parietal | 424 | 2.53 | −0.04 [−0.07, −0.02] |
| | | LH fusiform | 259 | 2.41 | −0.05 [−0.08, −0.02] |
| | | LH paracentral | 456 | 2.46 | −0.06 [−0.09, −0.03] |
| | | LH superior frontal & caudal middle frontal | 341 | 2.44 | −0.05 [−0.07, −0.02] |
| | | RH rostral middle frontal, superior frontal & medial orbitofrontal | 1646 | 2.42 | −0.05 [−0.08, −0.03] |
| | | RH superior frontal | 1878 | 2.43 | −0.07 [−0.10, −0.03] |
| | | RH paracentral, superior frontal & posterior cingulate | 1182 | 2.43 | −0.07 [−0.10, −0.04] |
| | | RH superior temporal & temporal pole | 925 | 2.54 | −0.10 [−0.15, −0.05] |
| | | RH superior frontal & posterior cingulate | 499 | 2.42 | −0.06 [−0.10, −0.03] |
| | | RH precentral, insula & postcentral | 542 | 2.39 | −0.05 [−0.07, −0.02] |
| | | RH caudal anterior cingulate & superior frontal | 407 | 2.44 | −0.06 [−0.09, −0.03] |
| | | RH superior temporal | 369 | 2.47 | −0.07 [−0.10, −0.03] |

***Note.*** These results are adjusted for age at neuroimaging assessment, sex, country of origin, education level, household income, BMI, and alcohol use before pregnancy. LH = left hemisphere, RH = right hemisphere

**Table S6. Vertex-wise analyses full results for area**

| Variable | Levels | Regions | Number of vertices | Mean area | Area mm [95% CI] |
|---|---|---|---|---|---|
| Smoking trajectories | Before pregnancy quitting vs non-smoking | LH middle temporal & inferior temporal | 875 | 0.59 | 0.04 [0.02, 0.06] |
| | | RH middle temporal & inferior temporal | 901 | 0.58 | 0.04 [0.02, 0.06] |
| Single time-point marker of smoking | Pack-years | LH fusiform & inferior temporal | 790 | 0.55 | -0.002 [-0.003, -0.001] |

***Note.*** These results are adjusted for age at neuroimaging assessment, sex, country of origin, education level, household income, BMI, and alcohol use before pregnancy. LH = left hemisphere, RH = right hemisphere

**Figure S1. Cortical thickness differences across smoking trajectories: brain regions showing overlap between persistent smoking, relapsing, and non-smoking**

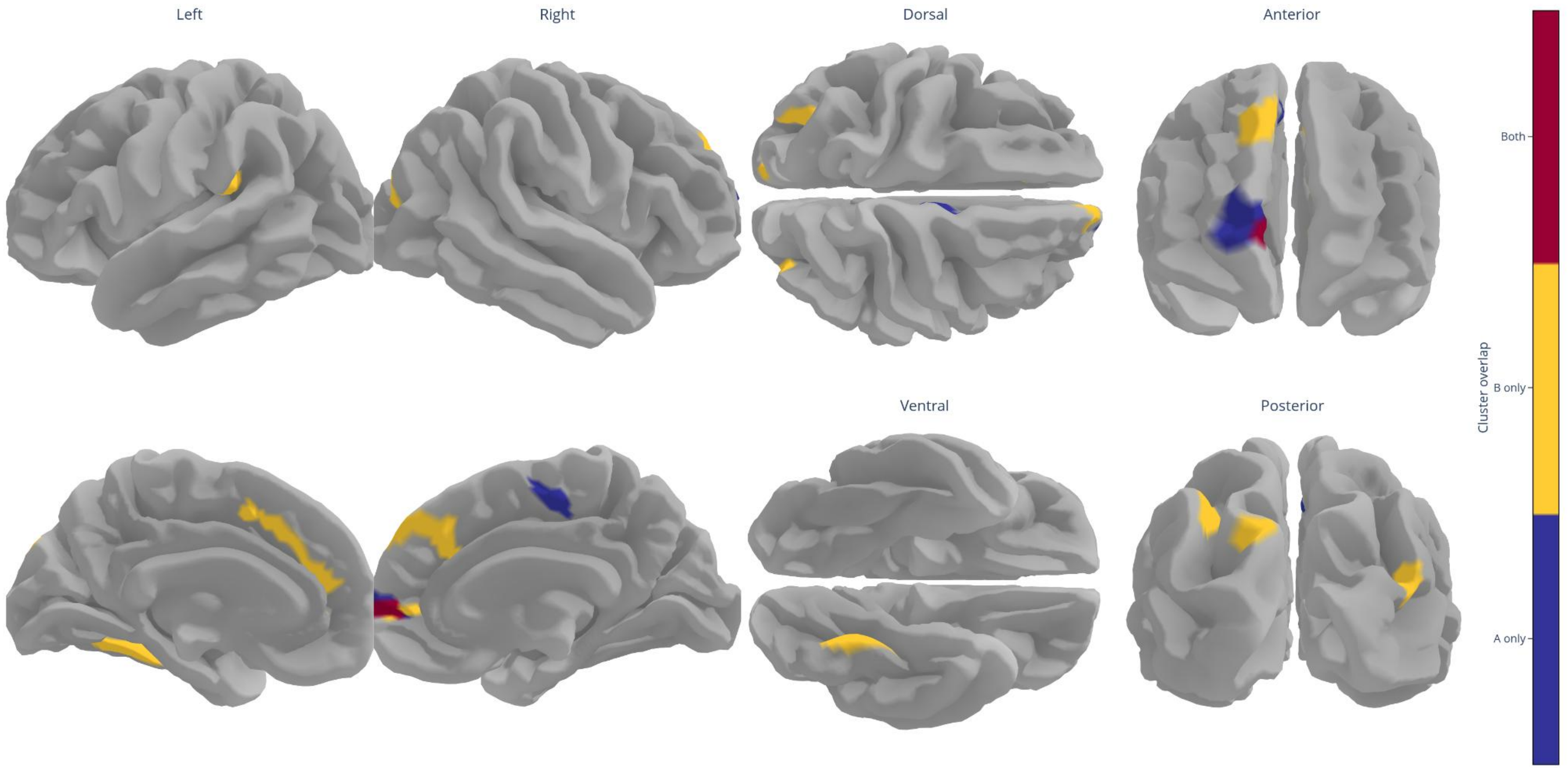


***Note.*** Yellow represents vertices with significant differences for persistent smoking (88.0%, 7042 vertices) vs non-smoking,, blue for relapsing (9.5%, 763 vertices) vs non-smoking, and red indicates overlap between these findings (2.5%, 199 vertices). These are results from the fully adjusted model, which accounts for age at neuroimaging assessment, sex, country of origin, education level, household income, BMI, and alcohol use before pregnancy. Vertex-wise analyses were cluster-wise corrected for multiple testing. Visualization was done with verywise (https://seredef.github.io/verywise/).

**Figure S2. Associations between single time-point markers of smoking and regional measures of cortical thickness and surface area**

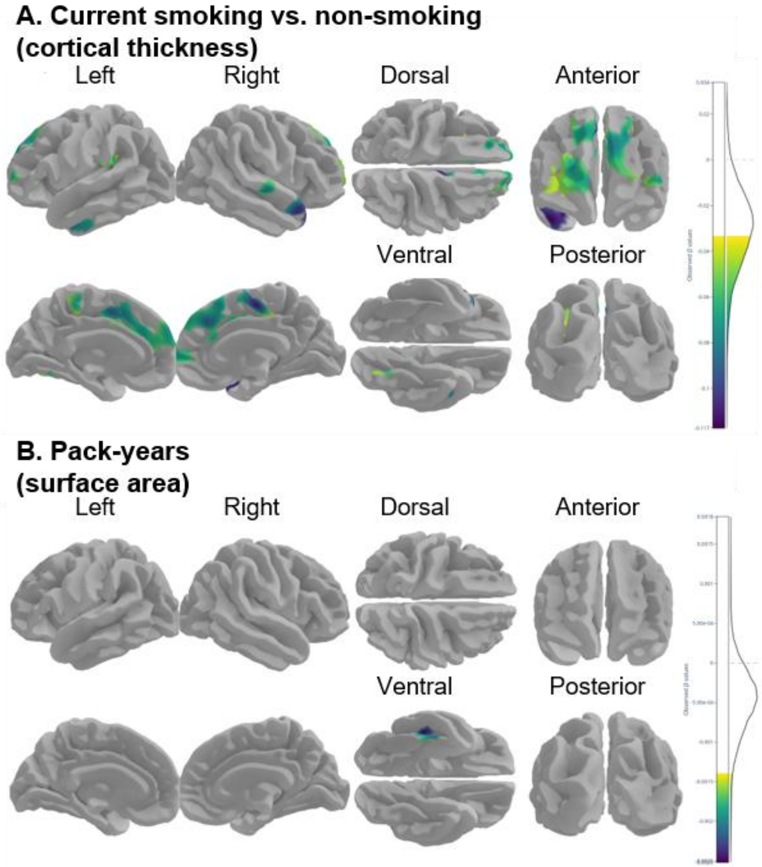


***Note.*** These are results from the fully adjusted model, which accounts for age at neuroimaging assessment, sex, country of origin, education level, household income, BMI, and alcohol use before pregnancy. Vertex-wise analyses were cluster-wise corrected for multiple testing. Visualization was done with verywise (https://seredef.github.io/verywise/).
